# Trends and Relationships in Opioid Prescribing Rates and Overdose Rates in the United States 2013-2019

**DOI:** 10.1101/2022.12.04.22283072

**Authors:** Luke F. Enthoven

## Abstract

The opioid epidemic began more than two decades ago and continues to increase in severity with the rate of overdose death increasing steadily. The epidemic was born in problematic claims about opioids and aggressive prescribing. After initial analysis of the first decade of the epidemic it became clear that actions would need to be taken to curtail overprescribing and many states passed legislation tracking and limiting opioid prescribing. Since then opioid prescribing has declined, but overdose has continued to increase at an alarming rate. This analysis is focused on evaluating more recent trends in opioid prescribing and overdose to see if the close positive correlation that existed between these values prior to 2010 holds true for the following decade. My findings indicate that between 2013 and 2019 that there is no relationship, or possibly a negative relationship, between opioid prescribing and overdose. Between 2013 and 2019 there was a 25% decrease in opioid prescribing rate yet a 96% increase in opioid overdose deaths. The close relationship that existed between prescribing and overdose appears to have diverged to the point where overdose increases as prescribing decreases. Linear regression on both national and state data revealed a significant negative relationship between overdose and prescribing for national data between 2013 and 2019 (R^2^ value of 0.84 and a p-value of 0.003) and no relationship between overdose and prescribing for state data between 2014 and 2019 (R^2^ value of 0.04 and a p-value of < 0.001). This questions the value of further efforts to reduce prescribing in preventing overdose and supports the hypothesis that restriction of prescription opioids can contribute to increased use of dangerous illicit opioids.

## Introduction

The modern opioid crisis has its roots in problematic prescribing practices during a period of aggressive introduction of new opioid medications and a push for increased awareness and treatment of patient pain. Opioid prescribing ramped up heavily from 1995 to 2010 with billions of prescriptions dispensed all over the United States during this period [1]. With this increased medical opioid availability came an increase in diversion and illicit use which led to addiction and physical dependence among both medical and non-medical users [2,3]. As opioid drugs became more popular and the market more saturated, opioid overdoses began to rise in the United States.

The first decade or so of the epidemic (1995-2009) was unique in that a majority of overdoses resulted from misuse of these new and widely available potent prescription opioids. These drugs were to blame for the almost four fold increase in overdose deaths in this time span, far outpacing any other group of illicitly used substances [4]. It is also relevant that during these years there was little to no change in rates of heroin use, and fentanyl was not yet widely available in illicit markets. Prescription opioids were clearly preferred by opioid users [5,6].

Due to the majority of opioid users using mainly prescription opioids prior to 2010, the availability of these drugs was highly linked to the rate of overdose. This relationship can most clearly be seen in the 2015 publication by Kolodny et. al. [7] which shows the close relationship between opioid sales, opioid overdose deaths, and opioid treatment admissions from 1999 to 2010. The authors suggest more conservative prescribing as one of many ways to reduce overdose mortality [7].

This same idea guided policy in response to the crisis. By the end of 2011, 49 states had passed legislation establishing some sort of prescription monitoring program. In 2015 the U.S Department of Health and Human Services called for “strong steps” to prevent addiction and overdose, including training, prescription monitoring programs, naloxone availability, and expanded use of medication-assisted treatment for addiction [8].

The goal of this investigation is to evaluate the more recent relationship between opioid prescribing and overdose using publicly available data from the National Institute on Drug Abuse (NIDA) [9], the Centers for Medicare and Medicaid Services (CMS) [10], and the Centers for Disease Control Wonder Databases (CDC) [11]. I assess if the correlation that was highlighted in Kolodny et. al. [7] is still what is observed when looking at the relationship between prescribing and overdose. The analysis also serves to evaluate the merit of continued efforts to curtail opioid prescribing in an attempt to reduce opioid mortality that are still popular among the medical community [12,13,14,15,16]

## Results

This investigation utilized data that is publicly available through NIDA, CMS, and the CDC. Overdose data was acquired through the NIDA and the CDC while opioid prescribing data was obtained from CMS. While CMS only provides data on prescribing under Medicare Part D, almost 50 million Americans are currently enrolled and therefore the prescribing rates provided by the CMS for Part D will be used to approximate national prescribing rates.

Prescribing rates and overdose rates were compared in multiple ways including nationally by year and by state by year. Data for years 2013 to 2019 was used, but not all analyses cover this whole span. CMS data showed the national opioid prescribing rate decreased 25% between 2013 and 2019 and an AMA release reported a 44.4% decrease in prescribing between 2011 and 2020 [17]. During this same period CDC data shows a 96% increase in the rate of overdose deaths. NIDA data showed the overdose rate to increase in every state from 2013-2019 except for Arkansas, Oklahoma, and Wyoming while CMS data showed a decrease in the prescription rate in every state. All three comparisons show either no relationship between opioid prescribing and overdose or a negative relationship between opioid prescribing and overdose. *Figure 1* and *Figure 2* use opioid overdose rate while *Figure 3* uses total overdose rate as state level opioid overdose data is not readily available. Due to such a large proportion of total overdoses involving opioids, total overdose rate is used as an approximation for opioid overdose rate in analysis using state level data in *Figure 3*. The percentage of overdoses involving opioids in the United States increased over time from 57% in 2013 to 70% in 2019 [9]. *Figure 1* shows a comparison between opioid prescribing and opioid overdose over time and highlights the increase in overdose rate and decrease in prescription rate that occurred between 2013 and 2019.

**Figure 1.**
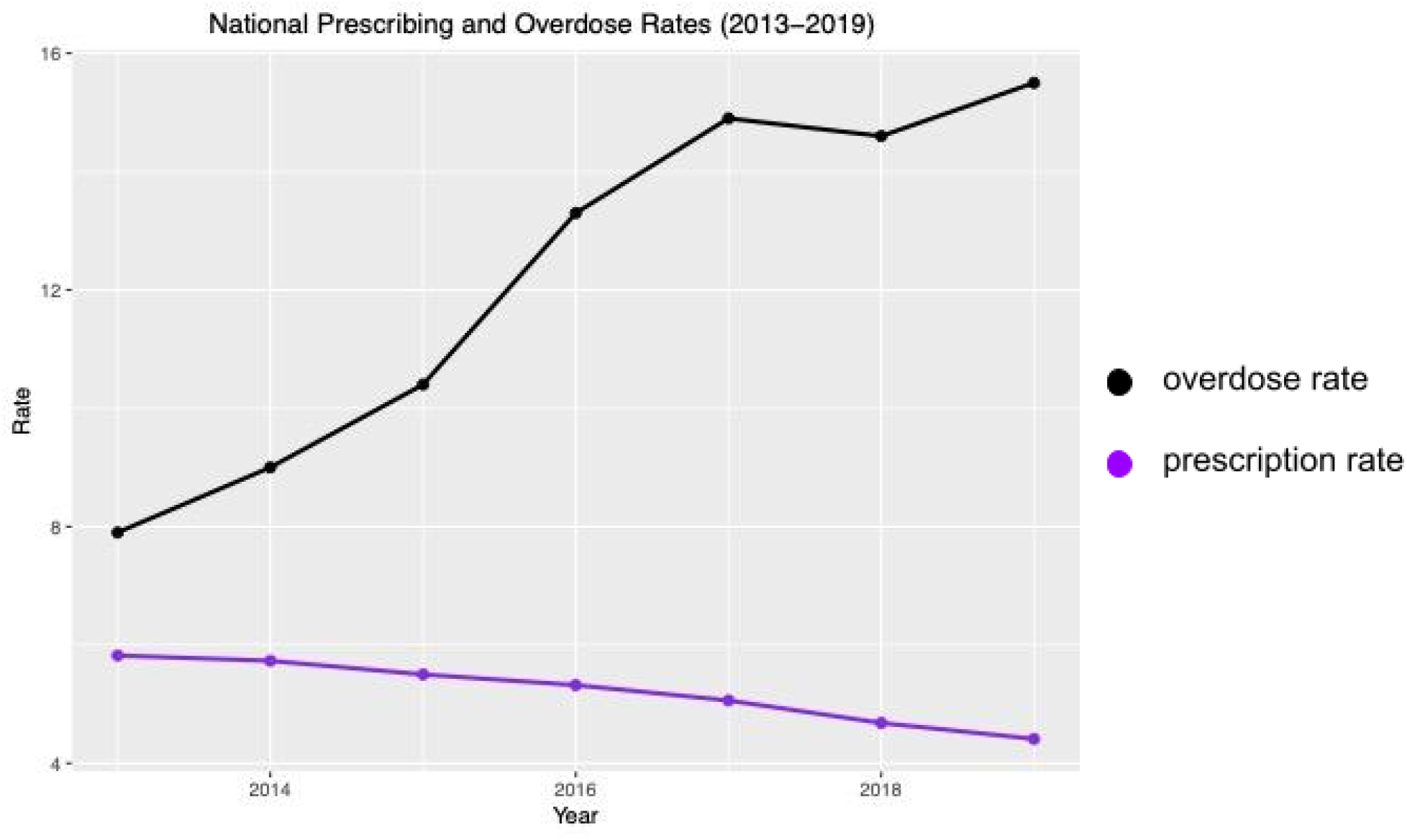
Opioid overdose rate is calculated as age-adjusted overdose deaths per 100,000 total population. Prescription rate is calculated as total Medicare Part D opioid claims divided by total Medicare Part D prescription claims. Data obtained from CMS and NIDA.

**Figure 2.**
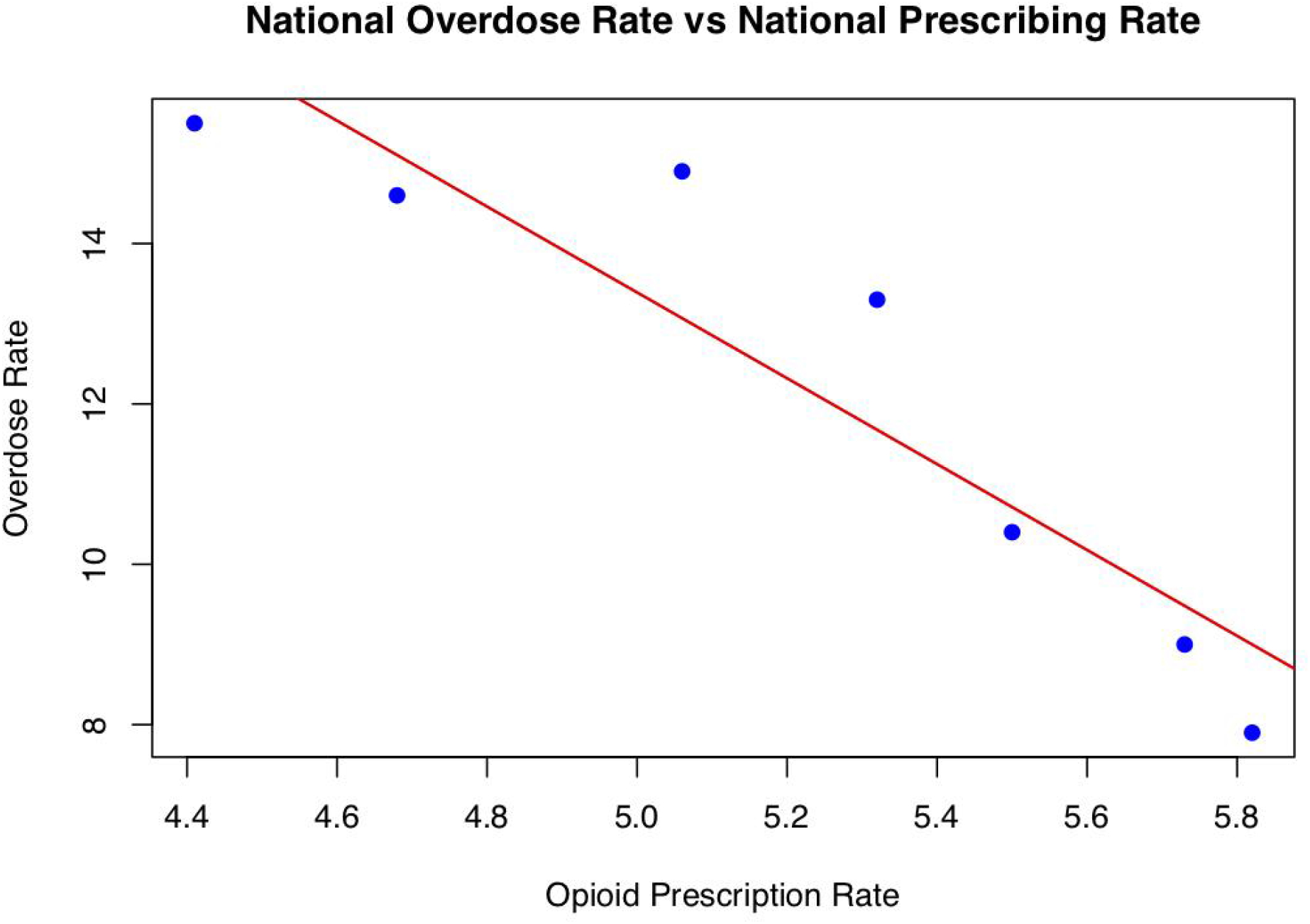
Opioid overdose rate is calculated as overdose deaths per 100,000 population. Prescription rate is calculated as total Medicare Part D opioid claims divided by total Medicare Part D prescription claims. Each data point represents the prescribing and overdose values for the United States at a given year between 2013 and 2019 for a total of 7 national snapshots. The red line is the result of a linear regression with an R^2^ value of 0.84 and a p-value of 0.003.

**Figure 3.**
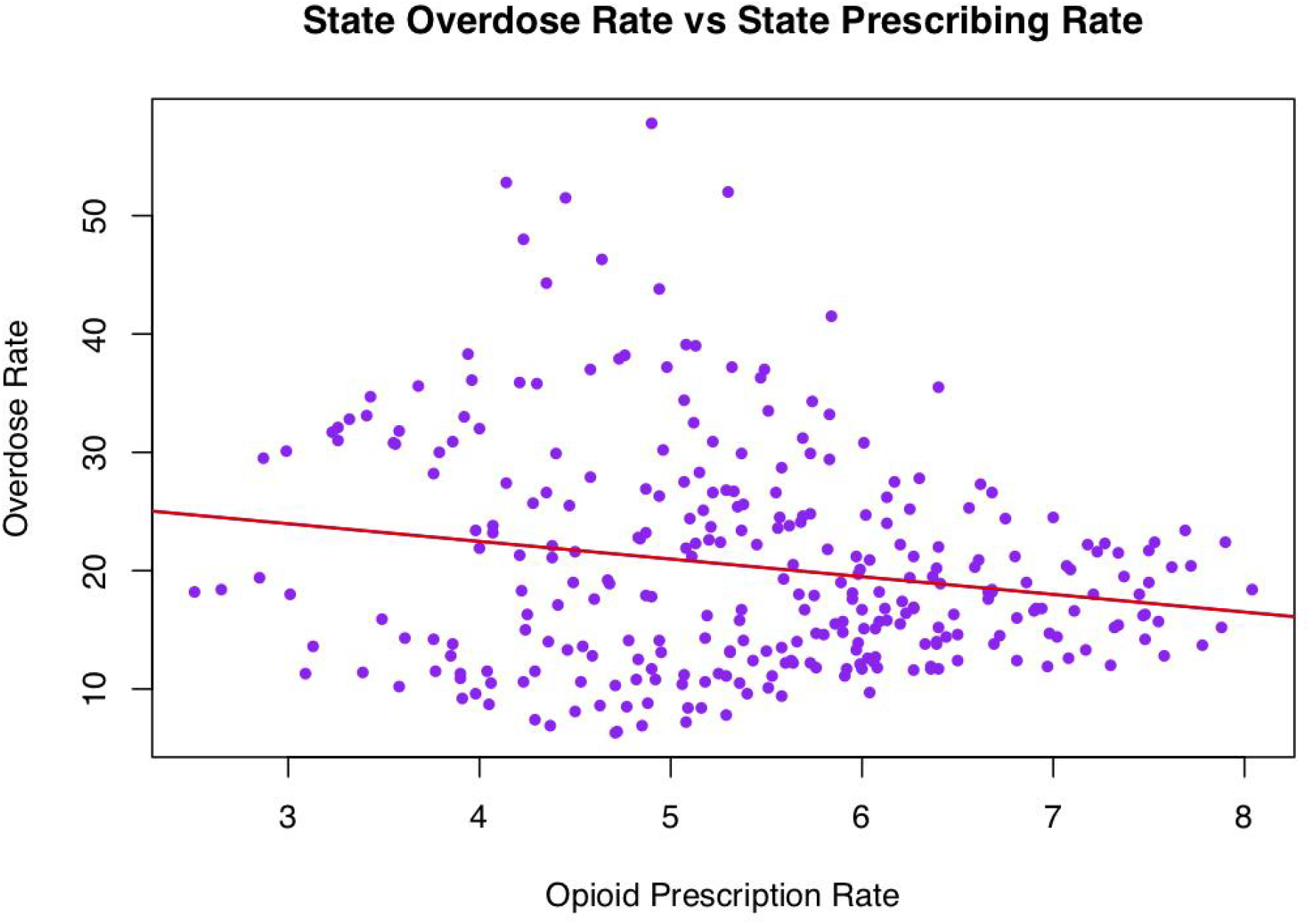
Overdose rate is calculated as overdose deaths per 100,000 population. Prescription rate is calculated as total Medicare Part D opioid claims divided by total Medicare Part D prescription claims. Each data point represents the prescribing and overdose values for a state at a given year between 2014 and 2019 for a total of 300 state snapshots. The red line is the result of a linear regression with an R^2^ value of 0.04 and a p-value of < 0.001. Overall overdose rate is used not opioid overdose rate specifically in this figure and analysis.

In order to test the direct relationship between prescribing and overdose, they were compared directly using state overdose and state prescribing data. The first of these is a reapplication of data from *Figure 1* that compares prescribing rate and overdose rate on the x and y axes respectively with each data point representing national overdose and prescribing in a given year between 2013 and 2019, *Figure 2*. This scatter was then subject to linear regression resulting in an R^2^ value of 0.84 and a p-value of 0.003. This indicates a significant negative relationship between prescribing rates and overdose rate in these limited annual national snapshots.

The trend shown in the regression for national annual data was strong, but limited by the small number of data points. This led to a comparison of prescribing rate and overdose rate with data points representing a single state in a single year. This takes away the temporal dimension of the data and simply investigates if a state’s prescription rate is related to its overdose rate based on state annual snapshots. The resulting scatter is upgraded from the 7 data points available in *Figure 2* to the 300 shown in *Figure 3*. This scatter was then subjected to a linear regression that gave an R^2^ value of 0.04 and a p-value of < 0.001. This would indicate that there is no relationship between overdose rate and prescribing rate in these annual state snapshots.

## Discussion

It is clear from both our data and similar wide and narrow scope analysis that efforts to reduce opioid prescribing in the United States have been extremely successful [18,19]. Prescribing rates are now as low as they were 20 years ago and continuing to trend downwards [20]. The goal of these changes, as previously stated, was to reduce diversion and abuse of prescription opioids which has also been very successful [21,22,23]. This success ultimately led to a decrease in deaths from prescription opioids in the United States, but appears to have had no effect, if not the opposite effect, on overall overdose mortality [23]. This supports other reports and analyses performed on this subject [17,24].

When the results of this analysis for 2013-2019 are compared to those found by Kolodny et. al., [7] it is clear that the close positive relationship that was once observed between opioid prescribing and overdose has diverged with more recent years showing no relationship or even a negative relationship. Despite the intended decrease in prescription misuse and prescription opioid overdose that inspired regulations, overdose mortality appears to be unaffected by efforts to limit prescription opioid availability [23]. Overall since implicating measures to restrict opioid prescriptions the rate of overdose has only increased in the United States with little indication of slowing [9]. These results continue to support the idea that those who abuse prescription opioids will not cease use when their drug of choice is not available, but instead will move to obtaining non pharmaceutical opioids from illicit sources. Such behavior increases risk of overdose and is reflected in the increased overdose risk for those whose opioid doses are reduced while on long term opioid therapy [25,26].

In the face of this evidence it does not seem efficacious to continue aiming efforts to solve the opioid epidemic at reducing prescribing. Prescribing has already been substantially reduced in the last decade and the changes have done little to impact trends in overdose mortality. Strategies such as patient education, harm reduction, accessible treatment, and physician training have been suggested as alternatives and are substantially supported in helping reduce overdose in the era of fentanyl [27,28,29].

## Limitations

This investigation also had its limitations. This was a very broad look at prescribing and overdose that did not dive deeper into understanding the many factors that influence both overdose and opioid prescribing. Data for study was not available for both metrics prior to 2013 and data was only available up until 2019. There has also been a rise in poly substance abuse. Our data for opioid overdose does not specify if multiple drugs were involved in mortality, making it more difficult to determine the exact role of opioids alone in contributing to overdose deaths.

## Methods

As previously stated all data used in this analysis is publicly available through NIDA, CMS, and the CDC. Data was obtained as CSV files that were used in R Studio in order to create figures and perform regression analysis [30]. In R Studio CDC overdose data and CMS prescribing data were combined in a data table for use in creating *Figure 1* and *Figure 2* using national data. NIDA overdose data and CMS prescribing data were combined for use in creating *Figure 3* using state data. *Figure 1* used the ggplot function in the ggplot2 package in order to create the plot [31]. *Figure 2* and *Figure 3* used the native R plot function to create the plots and display regression lines. Regression lines were calculated using the native R abline function. Lines were summarized in R to obtain R^2^ and p-values for the regression lines.

## Data Availability

All data produced are available online at

https://nida.nih.gov/research-topics/trends-statistics/overdose-death-rates

https://data.cms.gov/summary-statistics-on-use-and-payments/medicare-medicaid-opioid-prescribing-rates/medicare-part-d-opioid-prescribing-rates-by-geography

https://www.cdc.gov/nchs/pressroom/sosmap/drug_poisoning_mortality/drug_poisoning.htm

